# Liver Chemistries in COVID-19 Patients with Survival or Death: A Meta-Analysis

**DOI:** 10.1101/2020.04.26.20080580

**Authors:** Qing-Qing Xing, Xuan Dong, Yan-Dan Ren, Wei-Ming Chen, Dan-Yi Zeng, Yan-Yan Cai, Mei-Zhu Hong, Jin-Shui Pan

## Abstract

**Background and Aims:** Although abnormal liver chemistries are linked to higher risk of death related to coronavirus disease (COVID-19), liver manifestations may be diverse and even confused. Thus, we performed a meta-analysis of published liver manifestations and described the liver damage in COVID-19 patients with death or survival.

**Methods:** We searched PubMed, Google Scholar, medRxiv, bioRxiv, Cochrane Library, Embase, and three Chinese electronic databases through April 22, 2020. We analyzed pooled data on liver chemistries stratified by the main clinical outcome of COVID-19 using a fixed or random-effects model.

**Results:** In the meta-analysis of 18 studies, which included a total of 2,862 patients, the pooled mean alanine aminotransferase (ALT) was 30.9 IU/L in the COVID-19 patients with death and 26.3 IU/L in the COVID-19 patients discharged alive (p < 0.0001). The pooled mean aspartate aminotransferase (AST) level was 45.3 IU/L in the COVID-19 patients with death while 30.1 IU/L in the patients discharged alive (p < 0.0001). Compared with the discharged alive cases, the dead cases tended to have lower albumin levels but longer prothrombin time, and international standardized ratio.

**Conclusions:** In this meta-analysis, according to the main clinical outcome of COVID-19, we comprehensively described three patterns of liver impairment related to COVID-19, hepatocellular injury, cholestasis, and hepatocellular disfunction. Patients died from COVID-19 tend to have different liver chemistries from those are discharged alive. Close monitoring of liver chemistries provides an early warning against COVID-19 related death.

**Lay Summary:** Abnormal liver chemistries are linked to higher risk of death related to coronavirus disease (COVID-19). We performed a meta-analysis of 18 studies that included a total of 2,862 patients with COVID-19. We noted that patients died from COVID-19 tend to have different liver chemistries from those are discharged alive and close monitoring of liver chemistries provides early warning against COVID-19 related death.

## INTRODUCTION

Coronavirus disease (COVID-19) is caused by severe acute respiratory syndrome coronavirus 2 (SARS-CoV-2) infection, which is non-segmented positive-sense RNA viruses with envelope that belongings to the family Coronaviridae. Coronavirus widely distributes in humans and other mammals. In the past twenty years, coronavirus has caused several localized epidemics and even global pandemics, such as severe acute respiratory syndrome (SARS), Middle East Respiratory Syndrome (MERS), and the ongoing COVID-19. Worldwide, the spread and upward trend of COVID-19 have accelerated dramatically. According to the situation report released by the World Health Organization (WHO), as of April 25, 2020, 2,719,897 coronavirus disease (COVID-19) cases were confirmed globally with a fatality rate of 6.9% [1]. In response to the emerging threat, the WHO has declared a Public Health Emergency of International Concern on January 30, 2020, and further labeled as a pandemic on March 11, 2020. Compared with SARS and MERS, the case fatality rate of COVID-19 is relatively lower, which are 6.8% and 60% in SARS and MERS, respectively [2, 3]. Although the case fatality rate of COVID-19 is lower than those of SARS and MERS, the reproductive numbers of COVID-19 is even higher than of SARS [4]. Due to the huge number of confirmed cases, COVID-19 causes much more deaths than SARS or MERS.

Apart from lung injury, COVID-19 also leads to myocardial dysfunction, liver impairment, and acute kidney injury [5, 6]. Chronic hypertension and other cardiovascular comorbidities are frequent risk factors for COVID-19 related death [6, 7]. Other conditions can also contribute to death. According to the observation by Wu *et al* [7], coagulation dysfunction is linked to likelihood of death. Moreover, univariable logistic regression reveals the association between alanine aminotransferase (ALT), and total bilirubin (TBIL) and higher risk of death related to COVID-19 [8]. Another study by Fu *et al* [9] has shown the potential relationship between hypoproteinemia, cholestasis and higher fatality rate of the patients with COVID-19. Our meta-analysis also reveals the different patterns of abnormal liver chemistries between patients with severe and non-severe COVID-19 [10]. According to the clinical guideline proposed by American College of Gastroenterology, liver chemistries can be classified into three groups, in other words, hepatocellular injury-related indexes, including ALT and aspartate aminotransferase (AST); cholestatic injury-related indexes, comprised of alkaline phosphatase (AKP) and γ-glutamyltransferase (GGT); and hepatocellular function-related indexes such as albumin (ALB) level and prothrombin time (PT) [11]. In clinical practice, TBIL and direct bilirubin (DBIL), globulin (GLB) levels, and international standardized ratio (INR) are also assessed. Clinical manifestations of COVID-19 are diverse and even confusing. Comprehensive evaluation of the abnormalities of liver chemistries in patients with COVID-19 are rather few. Hence, the aim of this study was to provide a comprehensive view of liver test parameters in the COVID-19 patients with death or survival.

## METHODS

### Studies selection

The following databases were searched from December 1, 2019 to April 22, 2020, including PubMed, Google Scholar, medRxiv, bioRxiv, Embase, Cochrane Library, and three Chinese electronic databases (Chinese National Knowledge Infrastructure CQVIP, and Wanfang Data). “COVID-19”, “Coronavirus”, “SARS-CoV-2”, “2019-nCoV-2” or novel coronavirus were set as keywords for searching. The PRISMA guideline was implemented in the retrieval of potential studies[12]. Details of searching PubMed were listed in the Supplementary file. Endnote X9.3 (Thompson and Reuters, Philadelphia, Pennsylvania) was employed to manage the retrieved articles. Duplicated articles were removed.

### Selection criteria

Two authors (XD and QQX) independently determined the eligibility of the potential studies. Dissonance was arbitrated by the third author (JSP). The inclusion criteria were listed as the following: (1) study population: adult COVID-19 patients with death or survival; (2) study design: case report, case series, retrospective cohort study, prospective cohort study, randomized controlled trial, and case-control study; and (3) language: studies published in English or Chinese. The exclusion criteria were as follows: (1) pregnant women or pediatric patients; (2) patients who lack of nucleic acid data or serology evidence of SARS-CoV2 infection; (3) asymptomatic patients with SARS-CoV2 infection; (3) study design: commentary, editorial, review article, meta-analysis; (4) results: Studies that only reported the percentages of the indexes related to liver chemistries rather than the mean or average values of the corresponding indices, and studies that included only one arm (for example, only data of dead cases were reported without data from survival cases, or only data of survival cases were reported without data from dead cases) were also excluded.

### Data extraction

For the eligible articles, the following items: first author, study location, sample size, patient age and sex, and liver chemistry-related indexes such as TBIL, DBIL, ALT, AST, GGT, AKP, and ALB levels, were recorded. The main outcome (dead or survival) of COVID-19 was also recorded.

### Data analysis

R statistical platform (version 3.2.3, R Foundation for Statistical Computing) was used for statistical analysis. Continuous variables submitted to normal distribution were expressed as mean ± SD, while median (interquartile range [IQR]) was employed for the expression of those variables conformed to a skewed distribution. The method developed by Luo et al [13] was employed to estimate the sample mean and SD for the continuous outcomes from the studies that provided summary data of median, minimum, and maximum values. Related online tool is provided at http://www.math.hkbu.edu.hk/~tongt/papers/median2mean.html. The 95% confidence interval (CI) was presented in Forest plot. Heterogeneity among studies was detected by the Cochran Q test, with a p value of <0.10 indicating significant heterogeneity. The proportion of total variation among the studies, from which heterogeneity was derived, was measured by the *I*^2^ statistics. *I*^2^ values of <25%, 25–75%, and >75% represent low, moderate, and high heterogeneity, respectively [14]. Funnel plot was used to evaluated publication bias. A subgroup analysis was performed according to the main clinical outcome—death or survival.

## RESULTS

### Characteristics of the studies included in the meta-analysis

Selection process of potential studies was depicted in Figure 1. From the identified 1,841 studies, 18 studies were included in the meta-analysis. The characteristics of the enrolled studies are listed in Table 1 [6–8, 15–29]. Information including the study location, sample size, patient age and sex, clinical outcome, TBIL, DBIL, ALT, AST, GGT, AKP, ALB, GLB, PT, and INR was recorded. The mean ages of COVID-19 patients discharged alive and died were 56.4 and 68.7 years, respectively (Supplementary Figure 1). In the enrolled studies, male patients accounted for 55.3%. Among the studies that reported death cases, death case accounted for 31.6% (range, 11.7–61.5%) of the cases.

**Table 1.**
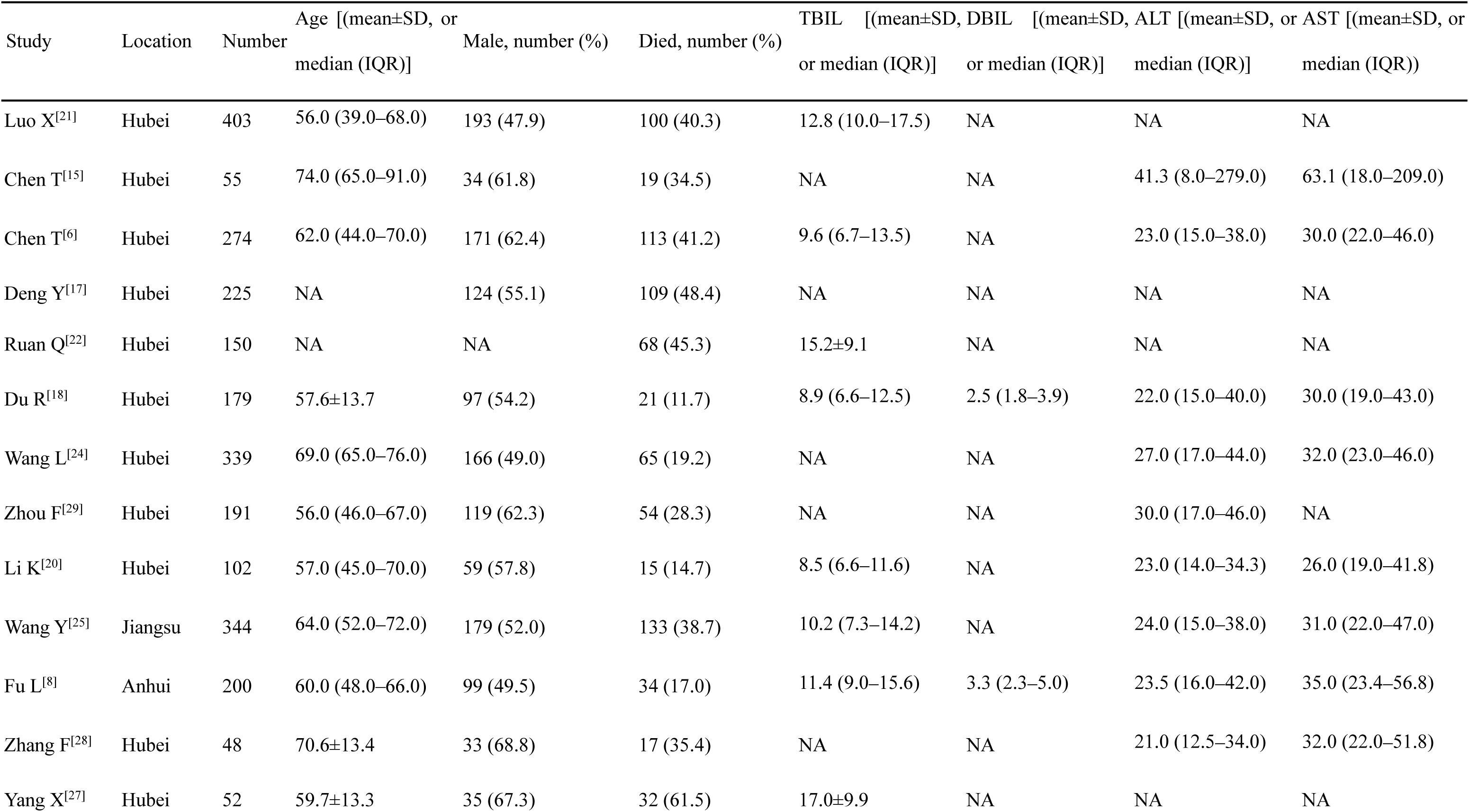
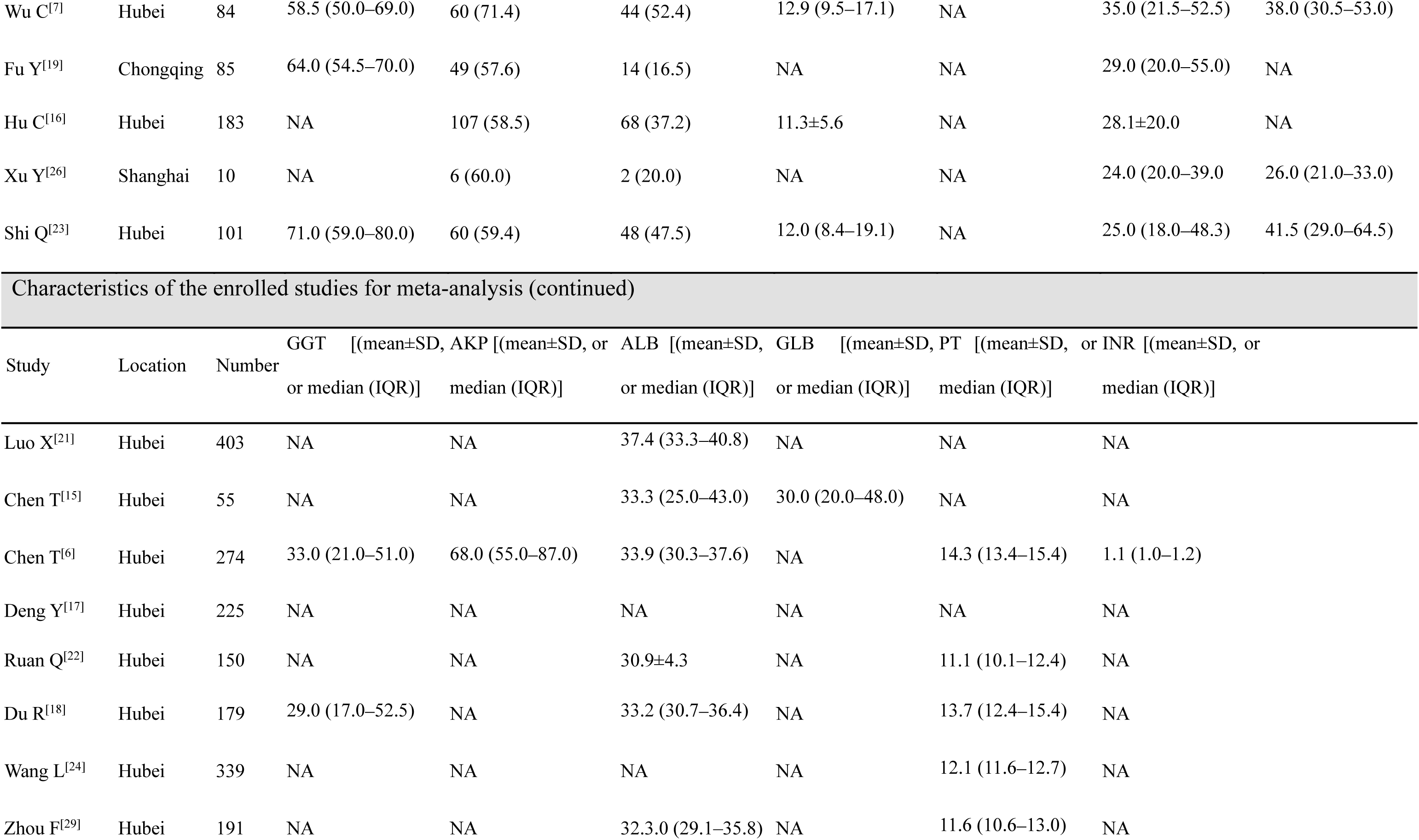
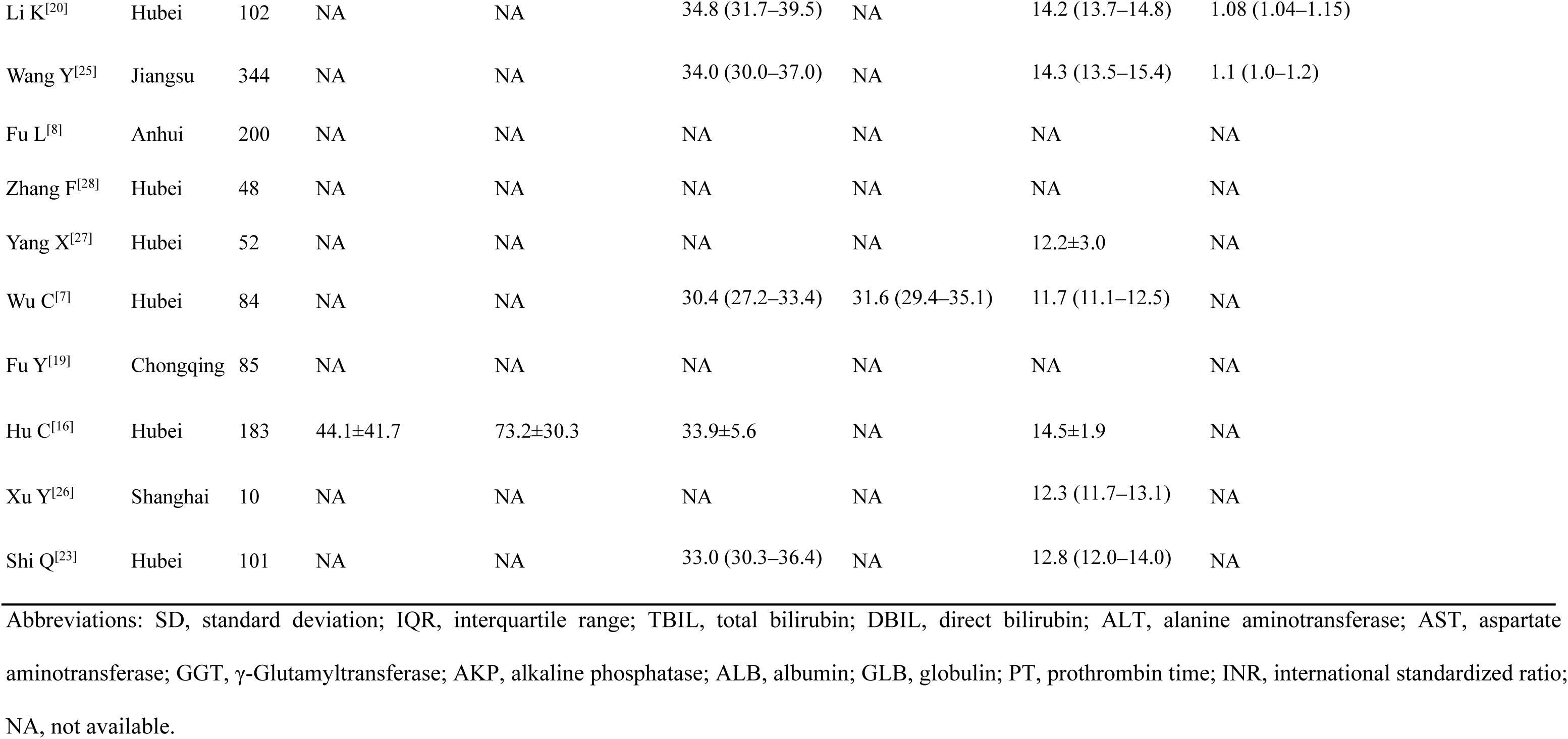
Characteristics of the enrolled studies for meta-analysis

| Study | Location | Number | Age [(mean±SD, or median (IQR))] | Male, number (%) | Died, number (%) | TBIL [(mean±SD, or median (IQR))] | DBIL [(mean±SD, or median (IQR))] | ALT [(mean±SD, or median (IQR))] | AST [(mean±SD, or median (IQR))] |
| --- | --- | --- | --- | --- | --- | --- | --- | --- | --- |
| Luo X<sup>[21]</sup> | Hubei | 403 | 56.0 (39.0–68.0) | 193 (47.9) | 100 (40.3) | 12.8 (10.0–17.5) | NA | NA | NA |
| Chen T<sup>[15]</sup> | Hubei | 55 | 74.0 (65.0–91.0) | 34 (61.8) | 19 (34.5) | NA | NA | 41.3 (8.0–279.0) | 63.1 (18.0–209.0) |
| Chen T<sup>[6]</sup> | Hubei | 274 | 62.0 (44.0–70.0) | 171 (62.4) | 113 (41.2) | 9.6 (6.7–13.5) | NA | 23.0 (15.0–38.0) | 30.0 (22.0–46.0) |
| Deng Y<sup>[17]</sup> | Hubei | 225 | NA | 124 (55.1) | 109 (48.4) | NA | NA | NA | NA |
| Ruan Q<sup>[22]</sup> | Hubei | 150 | NA | NA | 68 (45.3) | 15.2±9.1 | NA | NA | NA |
| Du R<sup>[18]</sup> | Hubei | 179 | 57.6±13.7 | 97 (54.2) | 21 (11.7) | 8.9 (6.6–12.5) | 2.5 (1.8–3.9) | 22.0 (15.0–40.0) | 30.0 (19.0–43.0) |
| Wang L<sup>[24]</sup> | Hubei | 339 | 69.0 (65.0–76.0) | 166 (49.0) | 65 (19.2) | NA | NA | 27.0 (17.0–44.0) | 32.0 (23.0–46.0) |
| Zhou F<sup>[29]</sup> | Hubei | 191 | 56.0 (46.0–67.0) | 119 (62.3) | 54 (28.3) | NA | NA | 30.0 (17.0–46.0) | NA |
| Li K<sup>[20]</sup> | Hubei | 102 | 57.0 (45.0–70.0) | 59 (57.8) | 15 (14.7) | 8.5 (6.6–11.6) | NA | 23.0 (14.0–34.3) | 26.0 (19.0–41.8) |
| Wang Y<sup>[25]</sup> | Jiangsu | 344 | 64.0 (52.0–72.0) | 179 (52.0) | 133 (38.7) | 10.2 (7.3–14.2) | NA | 24.0 (15.0–38.0) | 31.0 (22.0–47.0) |
| Fu L<sup>[8]</sup> | Anhui | 200 | 60.0 (48.0–66.0) | 99 (49.5) | 34 (17.0) | 11.4 (9.0–15.6) | 3.3 (2.3–5.0) | 23.5 (16.0–42.0) | 35.0 (23.4–56.8) |
| Zhang F<sup>[28]</sup> | Hubei | 48 | 70.6±13.4 | 33 (68.8) | 17 (35.4) | NA | NA | 21.0 (12.5–34.0) | 32.0 (22.0–51.8) |
| Yang X<sup>[27]</sup> | Hubei | 52 | 59.7±13.3 | 35 (67.3) | 32 (61.5) | 17.0±9.9 | NA | NA | NA |
| Wu C<sup>[7]</sup> | Hubei | 84 | 58.5 (50.0–69.0) | 60 (71.4) | 44 (52.4) | 12.9 (9.5–17.1) | NA | 35.0 (21.5–52.5) | 38.0 (30.5–53.0) |
| Fu Y<sup>[19]</sup> | Chongqing | 85 | 64.0 (54.5–70.0) | 49 (57.6) | 14 (16.5) | NA | NA | 29.0 (20.0–55.0) | NA |
| Hu C<sup>[16]</sup> | Hubei | 183 | NA | 107 (58.5) | 68 (37.2) | 11.3±5.6 | NA | 28.1±20.0 | NA |
| Xu Y<sup>[26]</sup> | Shanghai | 10 | NA | 6 (60.0) | 2 (20.0) | NA | NA | 24.0 (20.0–39.0) | 26.0 (21.0–33.0) |
| Shi Q<sup>[23]</sup> | Hubei | 101 | 71.0 (59.0–80.0) | 60 (59.4) | 48 (47.5) | 12.0 (8.4–19.1) | NA | 25.0 (18.0–48.3) | 41.5 (29.0–64.5) |

| Study | Location | Number | GGT [(mean±SD, or median (IQR))] | AKP [(mean±SD, or median (IQR))] | ALB [(mean±SD, or median (IQR))] | GLB [(mean±SD, or median (IQR))] | PT [(mean±SD, or median (IQR))] | INR [(mean±SD, or median (IQR))] |
| --- | --- | --- | --- | --- | --- | --- | --- | --- |
| Luo X<sup>[21]</sup> | Hubei | 403 | NA | NA | 37.4 (33.3–40.8) | NA | NA | NA |
| Chen T<sup>[15]</sup> | Hubei | 55 | NA | NA | 33.3 (25.0–43.0) | 30.0 (20.0–48.0) | NA | NA |
| Chen T<sup>[6]</sup> | Hubei | 274 | 33.0 (21.0–51.0) | 68.0 (55.0–87.0) | 33.9 (30.3–37.6) | NA | 14.3 (13.4–15.4) | 1.1 (1.0–1.2) |
| Deng Y<sup>[17]</sup> | Hubei | 225 | NA | NA | NA | NA | NA | NA |
| Ruan Q<sup>[22]</sup> | Hubei | 150 | NA | NA | 30.9±4.3 | NA | 11.1 (10.1–12.4) | NA |
| Du R<sup>[18]</sup> | Hubei | 179 | 29.0 (17.0–52.5) | NA | 33.2 (30.7–36.4) | NA | 13.7 (12.4–15.4) | NA |
| Wang L<sup>[24]</sup> | Hubei | 339 | NA | NA | NA | NA | 12.1 (11.6–12.7) | NA |
| Zhou F<sup>[29]</sup> | Hubei | 191 | NA | NA | 32.3.0 (29.1–35.8) | NA | 11.6 (10.6–13.0) | NA |

|  |  |  |  |  |  |  |  |  |
| --- | --- | --- | --- | --- | --- | --- | --- | --- |
| Li K<sup>[20]</sup> | Hubei | 102 | NA | NA | 34.8 (31.7–39.5) | NA | 14.2 (13.7–14.8) | 1.08 (1.04–1.15) |
| Wang Y<sup>[25]</sup> | Jiangsu | 344 | NA | NA | 34.0 (30.0–37.0) | NA | 14.3 (13.5–15.4) | 1.1 (1.0–1.2) |
| Fu L<sup>[8]</sup> | Anhui | 200 | NA | NA | NA | NA | NA | NA |
| Zhang F<sup>[28]</sup> | Hubei | 48 | NA | NA | NA | NA | NA | NA |
| Yang X<sup>[27]</sup> | Hubei | 52 | NA | NA | NA | NA | 12.2±3.0 | NA |
| Wu C<sup>[7]</sup> | Hubei | 84 | NA | NA | 30.4 (27.2–33.4) | 31.6 (29.4–35.1) | 11.7 (11.1–12.5) | NA |
| Fu Y<sup>[19]</sup> | Chongqing | 85 | NA | NA | NA | NA | NA | NA |
| Hu C<sup>[16]</sup> | Hubei | 183 | 44.1±41.7 | 73.2±30.3 | 33.9±5.6 | NA | 14.5±1.9 | NA |
| Xu Y<sup>[26]</sup> | Shanghai | 10 | NA | NA | NA | NA | 12.3 (11.7–13.1) | NA |
| Shi Q<sup>[23]</sup> | Hubei | 101 | NA | NA | 33.0 (30.3–36.4) | NA | 12.8 (12.0–14.0) | NA |

Abbreviations: SD, standard deviation; IQR, interquartile range; TBIL, total bilirubin; DBIL, direct bilirubin; ALT, alanine aminotransferase; AST, aspartate aminotransferase; GGT, γ-Glutamyltransferase; AKP, alkaline phosphatase; ALB, albumin; GLB, globulin; PT, prothrombin time; INR, international standardized ratio; NA, not available.

**Figure 1.**
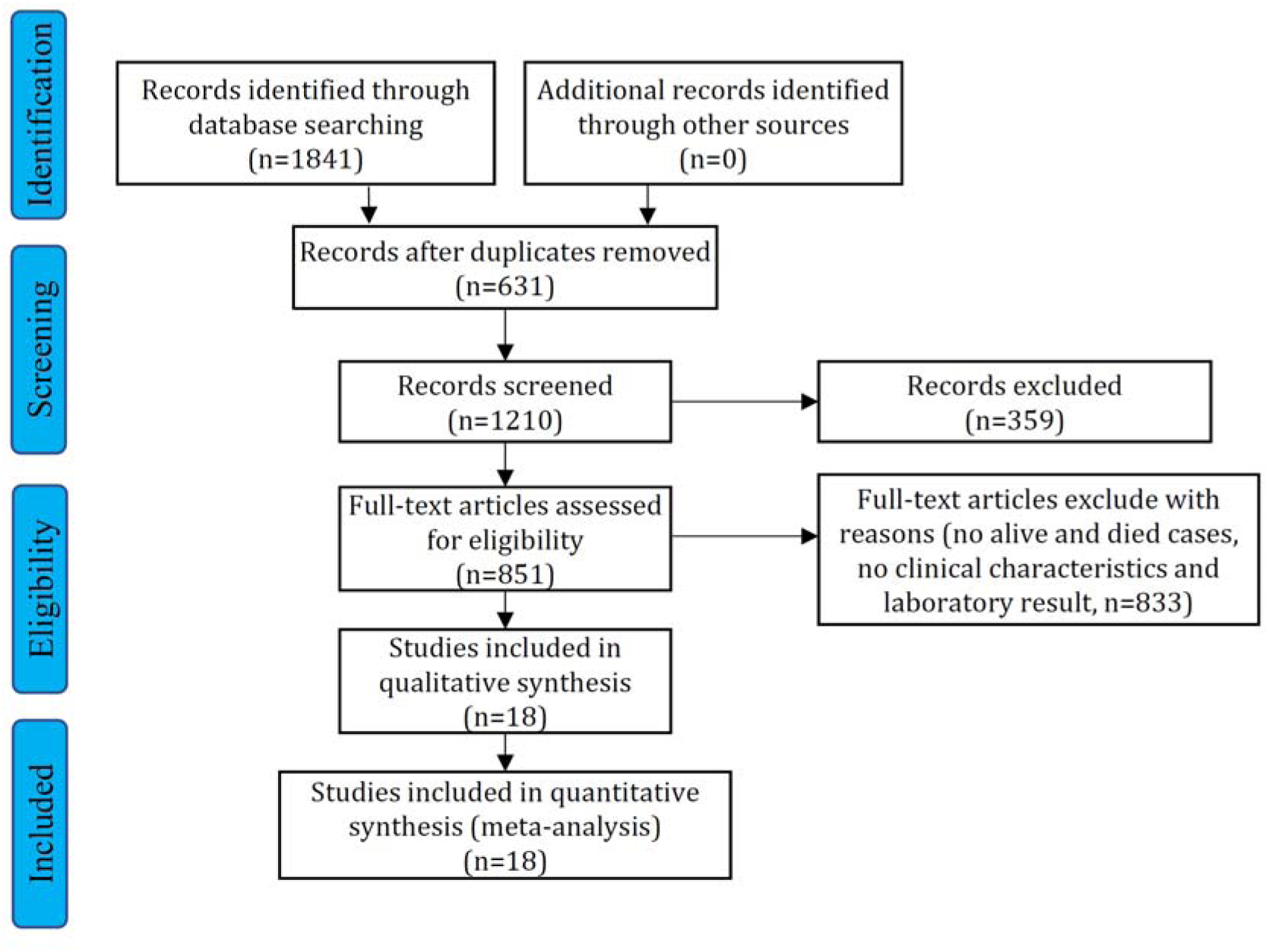
Study selection flow diagram If all the liver chemistry indexes were not reported, they were excluded from the meta-analysis. The studies that included only one arm (for example, only data of dead cases were reported without data from survival cases, or only data of survival cases were reported without data from dead cases) were also excluded.

### Hepatocellular injury-related abnormalities in liver chemistries

In the enrolled studies, 14 reported assays of ALT or AST in a total of 2,365 patients with COVID-19. All of these studies were from China. In the 18 enrolled studies, 14 (77.8%) were from Hubei Province, where Wuhan is located. The pooled mean ALT level was 30.9 IU/L in the COVID-19 patients with death and 26.3 IU/L in the COVID-19 patients discharged alive (95% CI: −6.1 to −2.9, p < 0.0001; Figure 2A), with moderate heterogeneity among the studies (*I*^2^ = 42%, p = 0.05). Similarly, the pooled mean AST level was 45.3 IU/L in the COVID-19 patients with death and 30.1 IU/L in the patients discharged alive (95% CI: −18.3 to −8.6, p < 0.0001; Figure 2B). Significant heterogeneity was observed for the AST levels among the studies (*I*^2^ = 74%, p < 0.01), which was significantly higher than that of the ALT levels (*I*^2^ = 42%, p = 0.05). Potential publication bias existed in ALT/AST was evaluated using a funnel plot (Supplementary Figure 2). In the patients with COVID-19, the mean AST level tended to be higher than the mean ALT level in both the died and discharged alive groups. Moreover, the gap between the AST and ALT levels (45.3 and 29.8 IU/L, respectively) was even more significant in the group discharged alive (Figure 3). Evaluation of potential publication bias was also presented in Supplementary Figure 3.

**Figure 2.**
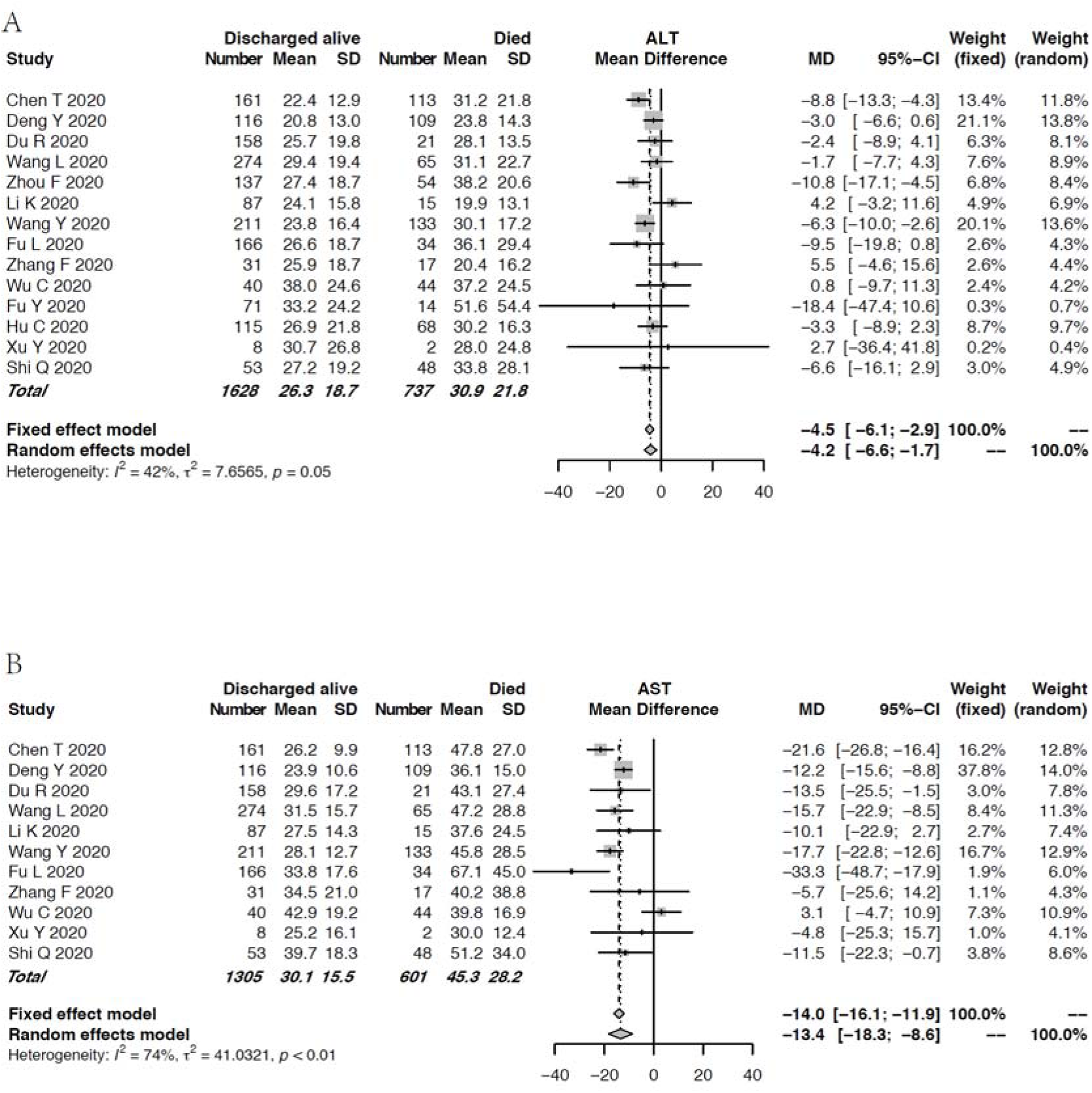
Forest plot of the association between serum ALT/AST level and the main clinical outcomes—died or discharged alive. Pooled levels of (A) ALT and (B) AST in the patients with COVID-19 stratified by clinical outcome. Abbreviations: ALT, alanine aminotransferase; AST, aspartate aminotransferase.

**Figure 3.**
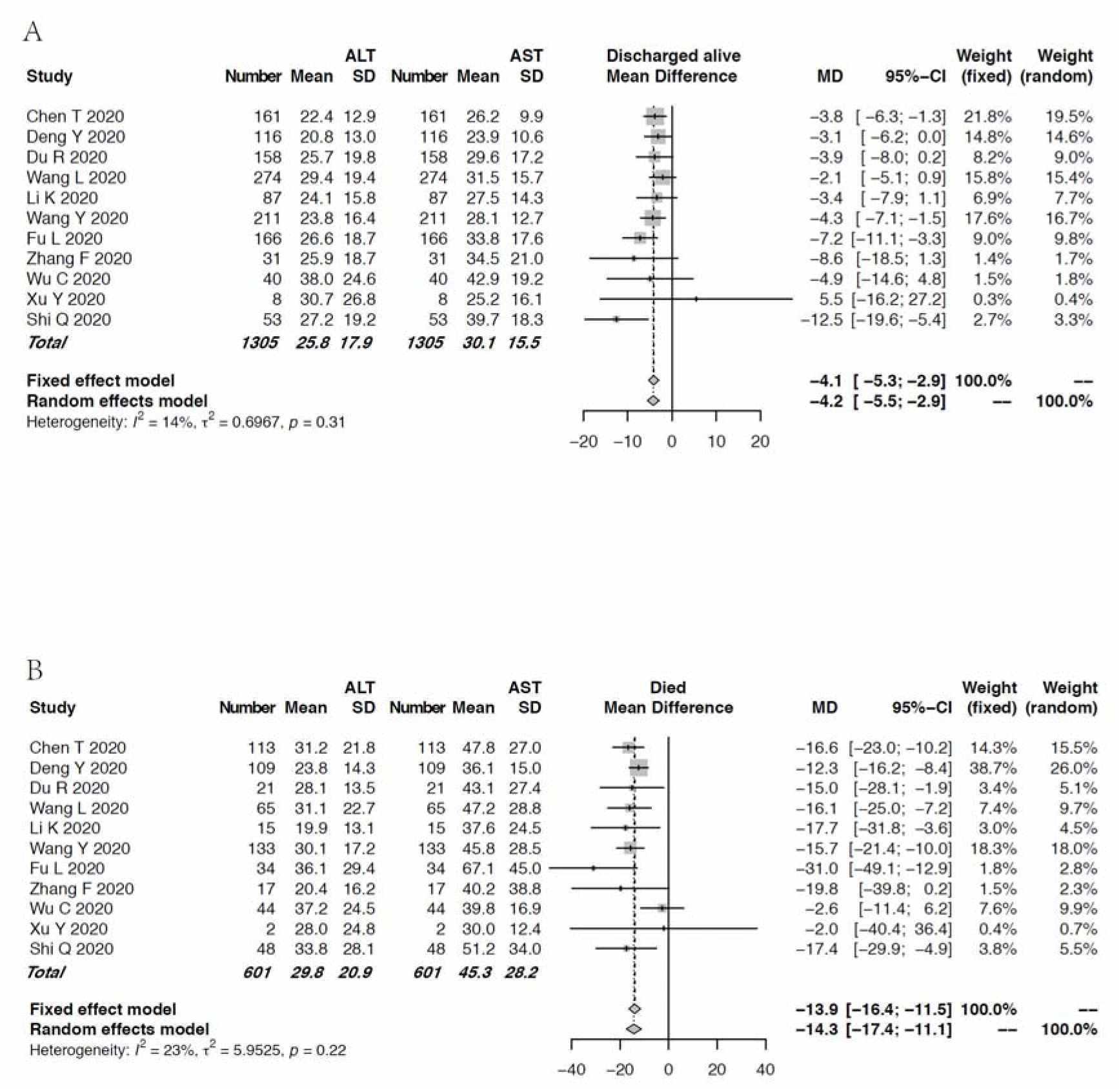
Forest plot for the comparison of ALT and AST levels in the patients with COVID-19 stratified by clinical outcome A, Forest plot for the comparison of ALT and AST levels in the group discharged alive. B, Forest plot for the comparison of ALT and AST levels in the group with death. Abbreviations: ALT, alanine aminotransferase; AST, aspartate aminotransferase.

### Cholestasis-related abnormalities in liver chemistries

We cannot enroll enough studies to analysis cholestasis-related indexes such as AKP, and DBIL levels, since rather few studies focused on these indexes. Of the enrolled studies for the meta-analysis, 3 reported GGT assays and 11 studies reported TBIL measurements. The pooled mean GGT level was 46.3 IU/L in the group with death and 34.3 IU/L in the group discharged alive (Figure 4A). The pooled mean TBIL level in the group with death was slightly higher than that in the group discharged alive. However, the mean TBIL levels remained within the normal range in both groups (Figure 4B). For the TBIL levels, moderate heterogeneity was observed among the studies (*I*^2^ = 36%, p = 0.11). Funnel plot for the TBIL levels was shown in Supplementary Figure 4.

**Figure 4.**
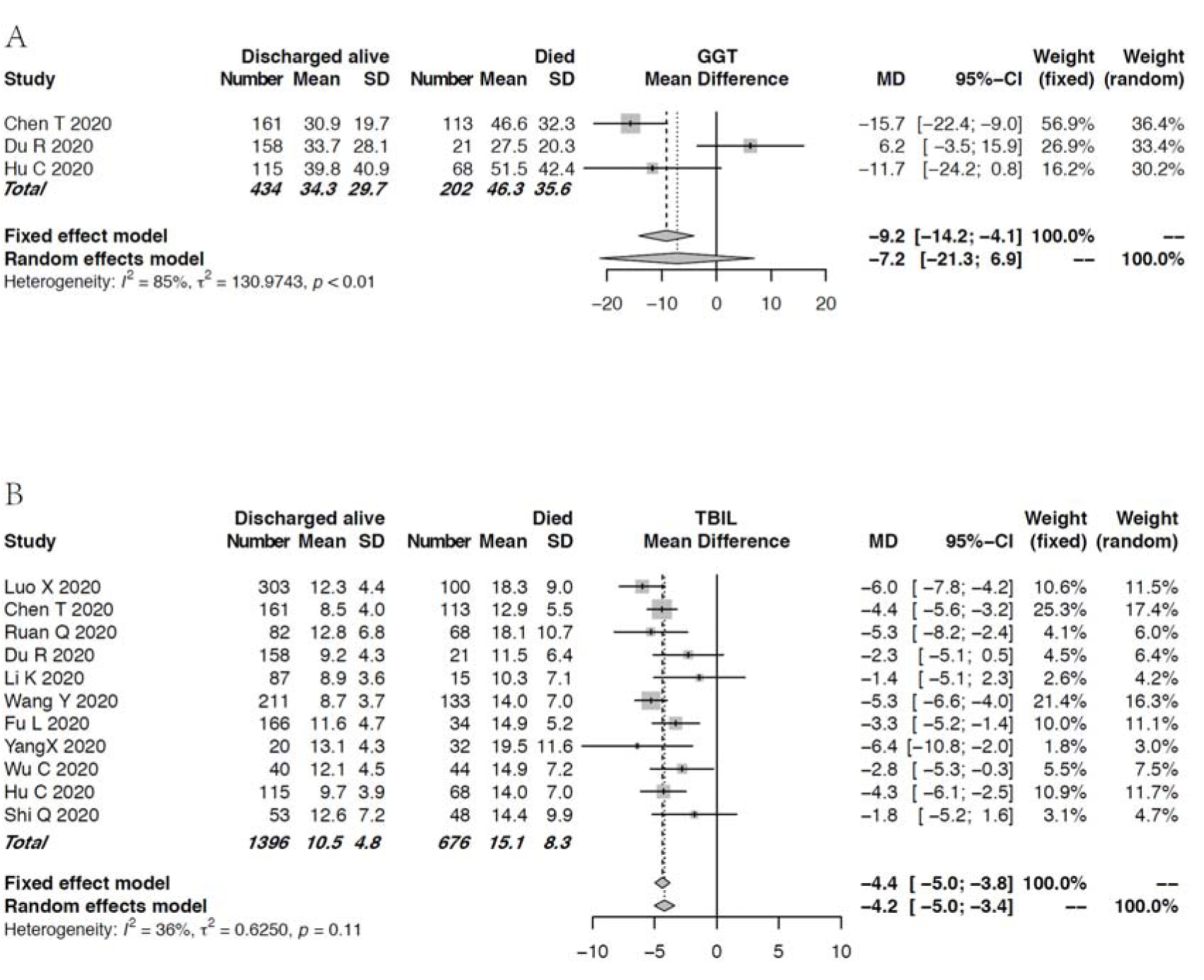
Forest plot for the association of the cholestasis-related indexes and clinical outcome. Pooled levels of (A) GGT, and (B) TBIL in the patients with COVID-19. Abbreviations: GGT, γ-Glutamyltransferase; TBIL, total bilirubin.

### Hepatocellular function-related abnormalities in liver chemistries

Eleven studies compared the mean ALB levels according to the main clinical outcome of COVID-19, between 683 and 1,383 died and discharged alive cases, respectively (Figure 5A). High heterogeneity was observed among the studies (*I*^2^ = 80%, p < 0.01). The mean ALB level in the patients with death was significantly lower than that in the patients discharged alive. There were significant differences in the coagulation-related indexes such as PT and INR between the two groups. The patients in the group with dead had longer PT or higher INR (Figure 5B, 5C). Evaluation of publication bias related to ALB level and PT was shown in Supplementary Figure 5.

**Figure 5.**
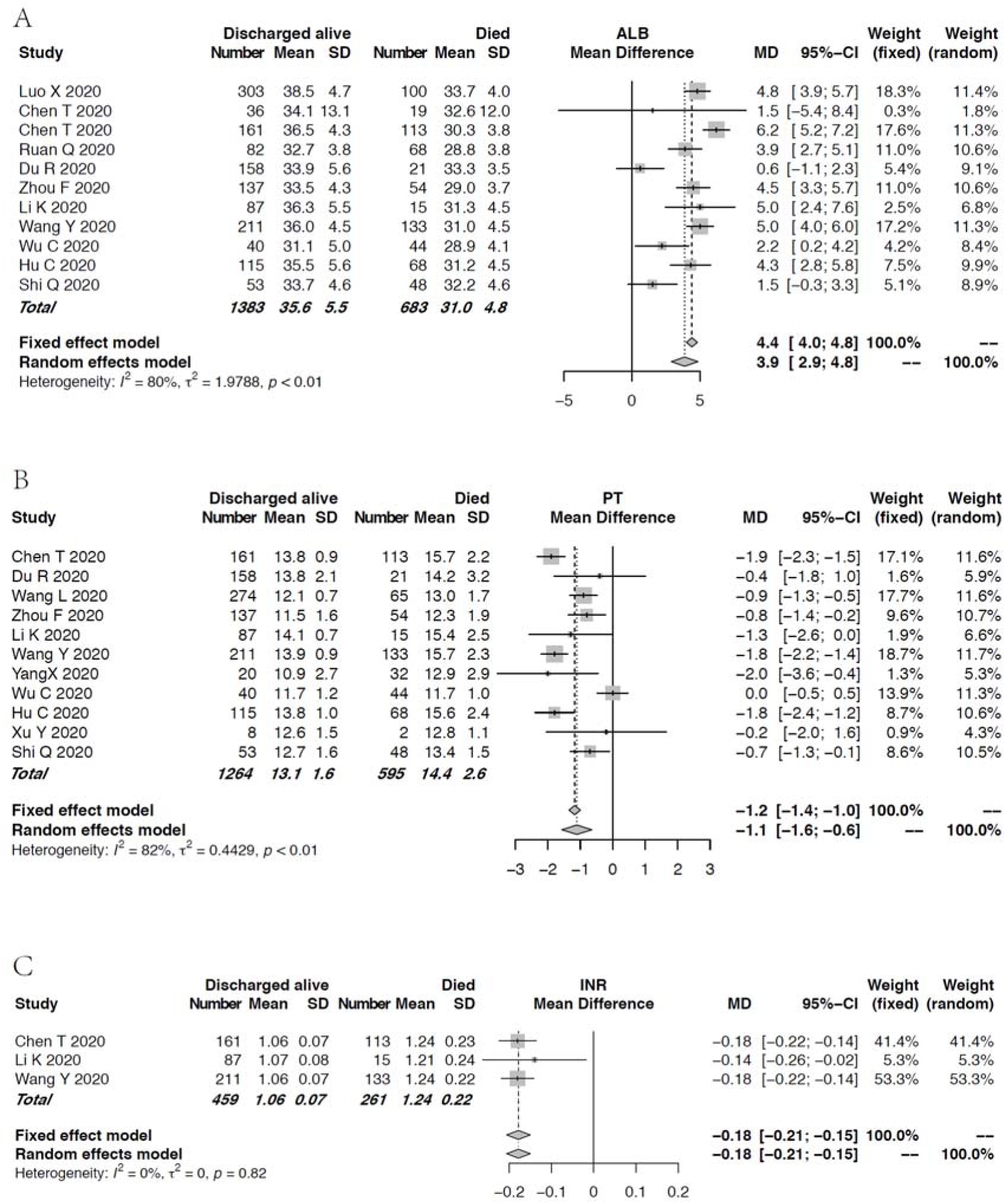
Forest plot for the association of the synthetic function-related indexes and clinical outcome Pooled (A) ALB levels, (B) PTs, and (C) INRs in the patients with COVID-19. Abbreviations: ALB, albumin; PT, prothrombin time; INR, international standardized ratio.

## DISCUSSION

In this meta-analysis, 18 studies, all from China, consisted a total of 3,025 patients with COVID-19 were enrolled. Three patterns of liver impairment, namely hepatocellular injury, cholestasis, and hepatocellular dysfunction, can develop in proportion of patients with COVID-19, especially in the patients who eventually died. Briefly, the patients who died of COVID-19 tended to have higher baseline ALT/AST, TBIL, INR, and prolonged PT while lower ALB than the survival cases. The patients with COVID-19 usually had higher AST levels than ALT levels, especially in the non-survival. There was a tendency of the dead cases to arise in the elderly. Should be noted, the pooled percentage of death cases accounted up 31.6%, which was calculated according to the proportion of death cases in each enrolled selected study. It does not intend to mean that the percentage of death cases in the whole population reaches that high. Several reasons that may lead to the higher percentages of death cases in the enrolled study than those in the whole population. First, since the patients in the enrolled studies are all hospitalized or even critically ill patients, it sounds reasonable that the percentage of death case is higher than that in the whole population. Second, the composition difference exists in the enrolled studies caused by the selection of the reporter. Similar to our previous study, most studies focus on ALT/AST rather than cholestasis-related indexes such as AKP and GGT levels [10]. In other words, the actual prevalence of abnormal liver chemistries could be underestimated in the studies reporting clinical features of COVID-19. Thus, the role of liver chemistries was compromised in disease monitoring and early warning against potential death cases.

The possible mechanisms of COVID-19 related liver injury include the direct damage of SARS-CoV-2 to the liver, and the secondary liver injury caused by stress and systemic inflammatory response, liver ischemia or hypoxia, exacerbation of underlying liver disease, drug-induced liver injury [30]. In addition, for HBsAg positive patients, the following events can also lead to liver injury: sudden cessation of ongoing antiviral treatment in the patients with chronic hepatitis B patients, which may result in sharp deterioration of liver chemistries or acute-on-chronic liver failure; application of high dose corticosteroids without antiviral therapy, which may bring about activate replication of HBV. Clinically, secondary liver injury is far more frequent than direct liver injury [31].

The most frequent complications of deceased patients caused by COVID-19 are the following: acute respiratory distress syndrome, sepsis, acute cardiac injury, type I respiratory failure, followed by heart failure [6]. The percentage of developing acute liver injury is 9% in deceased patients whereas is only 2% in recovered patients [6]. Investigation by Deng et al [17] also indicated the high frequency of developing acute respiratory distress syndrome (89.9%), acute cardiac injury (59.6%), acute kidney injury (18.3%) in the patients with death. Sepsis or liver ischemia caused by acute respiratory distress syndrome or heart failure can further bring about abnormalities of liver chemistries or even disordered coagulopathy.

On the whole, the alteration of liver chemistries between the survival group and non-survival group is similar to that between the severe group and the non-severe group, shown in our recent study [10], although there are some differences. Similar to the age difference between severe and non-severe group, our analysis also found that the average age of the non-survival group was significantly higher than that of survival group. The average ALT and AST in the dead group were also higher than those in the non-survival group. Moreover, both in the non-survival group and survival group, the average AST was higher than ALT, which was also similar to our previous study [10]. GGT in the non-survival group was higher than that in the survival group. However, there was no statistical significance. This may be due to the under-reported on GGT. TBIL in the non-survival group was also higher than that in the survival group whereas the average values of TBIL in both groups were all within the normal range. ALB in the non-survival group was significantly lower than that in the survival group, which was in accordance with our study between severe and non-severe group [10]. In the non-survival group, PT and INR were significantly longer than those in the survival group, which was even more remarkable than the difference we previously observed between the severe group and non-severe group. This suggests that the coagulation function of non-survival group is significantly worse than that of survival group. Since almost all of coagulation factors are synthesized in the liver, it can be speculative that the hepatocellular disfunction in the non-survival group was exacerbated under the influence of multiple factors such as hypoxemia, inflammation storm, infection. In fact, severe sepsis is almost invariably associated with systemic activation of coagulation [32, 33].

In this study, we found that the abnormalities of liver chemistries in the non-survival group was worse than that in the survival group, which suggested that we should be alert to the risk of death for the patients with significantly abnormal liver chemistries on admission. Dramatical hypoalbuminemia usually developed in the non-survival group. Thus, solid nutrition support is necessary. In addition, if there is obvious coagulation dysfunction on admission, we should be aware of the risk of severe case or even death. It is necessary to correct the coagulation disfunction as early as possible to prevent the further deterioration of the condition.

Several substantial merits can be seen in this study. First, the literatures focused on COVID-19 related abnormal liver chemistries are rapidly evolving and sometimes confusing. This meta-analysis comprehensively summarized the related literatures according to the main clinical outcome—death or survival. The extensive coverage of over 2,800 studies allowed a more precise evaluation of the abnormalities of liver chemistries in the patients with COVID-19. Our analysis revealed that abnormal liver chemistries were associated with a worse clinical outcome and even death, which highlights the importance of a more extensive monitoring on liver chemistries for the purposes of both diagnosis and prognosis. Second, three patterns of liver impairment, namely hepatocellular injury, cholestasis, and hepatocellular dysfunction were all extensively covered in this analysis. However, cholestasis-related indices such as AKP and GGT tended to be inadvertently ignored, which is even more significant than that in our previous study [10]. We also compared hepatocellular dysfunction between the survival and non-survival cases. The alarmingly high prevalence of hypoalbuminemia and coagulation dysfunction in the non-survival group requires vigilance. Third, we also covered the eligible studies preprinted in medRxiv, which ensured that our meta-analysis has a clear leading position. However, our study has a few limitations. As mentioned earlier, cholestasis-related indices were remarkably under-reported, which hindered us from getting more precise pooled data. Second, all the enrolled studies came from mainland China, which restricted a more precise estimation of abnormal liver chemistries in the context of diverse other ethnics. However, this conversely helps to abate the heterogeneity caused by the physiological differences.

## CONCLUSION

In this analysis, we comprehensively described three patterns of liver impairment related to COVID-19 including hepatocellular injury, cholestasis, and hepatocellular dysfunction, according to the main clinical outcome of COVID-19. Patients with abnormal liver chemistries are at higher risk of worse outcome. Close monitoring on liver chemistries, especially coagulation-related indexes, is beneficial to early warning against worse outcome.

## Data Availability

Not available.

## Acknowledgments

**Financial support:** This work was supported by the National Natural Science Foundation of China No. 81871645 (JSP). The funding source did not have any role in the design and conduct of the study; collection, management, analysis, and interpretation of the data; preparation, review, or approval of the manuscript; and decision to submit the manuscript for publication.

## Guarantor of the article

Professor Jin-Shui Pan

## Specific author contributions

JSP and MZH were involved with the study conceptualization and design; analysis and interpretation of data; drafting of the manuscript; and approval of the final version of the manuscript; XD, DYZ, YYC, WMC, QQX, and YDR were involved in data retrieval.

## REFERENCES

[1] World Health Organization. Coronavirus disease 2019 (COVID-19) Situation Report–96. World Health Organization; 25 April 2020.

[2] Donnelly CA, Ghani AC, Leung GM, Hedley AJ, Fraser C, Riley S, et al. Epidemiological determinants of spread of causal agent of severe acute respiratory syndrome in Hong Kong. Lancet 2003;361:1761–1766.

[3] Assiri A, Al-Tawfiq JA, Al-Rabeeah AA, Al-Rabiah FA, Al-Hajjar S, Al-Barrak A, et al. Epidemiological, demographic, and clinical characteristics of 47 cases of Middle East respiratory syndrome coronavirus disease from Saudi Arabia: a descriptive study. Lancet Infect Dis 2013;13:752–761.

[4] Liu Y, Gayle AA, Wilder-Smith A, Rocklov J. The reproductive number of COVID19 is higher compared to SARS coronavirus. J Travel Med 2020;27.

[5] Guan WJ, Ni ZY, Hu Y, Liang WH, Ou CQ, He JX, et al. Clinical Characteristics of Coronavirus Disease 2019 in China. N Engl J Med 2020.

[6] Chen T, Wu D, Chen H, Yan W, Yang D, Chen G, et al. Clinical characteristics of 113 deceased patients with coronavirus disease 2019: retrospective study. BMJ 2020;368:m1091.

[7] Wu C, Chen X, Cai Y, Xia J, Zhou X, Xu S, et al. Risk Factors Associated With Acute Respiratory Distress Syndrome and Death in Patients With Coronavirus Disease 2019 Pneumonia in Wuhan, China. JAMA Intern Med 2020.

[8] Fu L, Fei J, Xiang H-X, Xiang Y, Tan Z-X, Li M-D, et al. Influence factors of death risk among COVID-19 patients in Wuhan, China: a hospital-based case-cohort study. 2020:2020.2003.2013.20035329.

[9] Fu L, Fei J, Xu S, Xiang HX, Xiang Y, Tan ZX, et al. Acute liver injury and its association with death risk of patients with COVID-19: a hospital-based prospective case-cohort study. medRxiv 2020:2020.2004.2002.20050997.

[10] Dong X, Zeng DY, Cai YY, Chen WM, Xing QQ, Ren YD, et al. Liver Chemistries in Patients with Severe or Non-severe COVID-19: A Meta-Analysis (unpublished data). 2020.

[11] Kwo PY, Cohen SM, Lim JK. ACG Clinical Guideline: Evaluation of Abnormal Liver Chemistries. Am J Gastroenterol 2017;112:18–35.

[12] Liberati A, Altman DG, Tetzlaff J, Mulrow C, Gotzsche PC, Ioannidis JP, et al. The PRISMA statement for reporting systematic reviews and meta-analyses of studies that evaluate healthcare interventions: explanation and elaboration. BMJ 2009;339:b2700.

[13] Luo D, Wan X, Liu J, Tong T. How to estimate the sample mean and standard deviation from the sample size, median, extremes or quartiles? Chinese Journal of Evidence-Based Medicine 2017;17:1350–1356.

[14] Higgins JP, Thompson SG. Quantifying heterogeneity in a meta-analysis. Stat Med 2002;21:1539–1558.

[15] Chen T, Dai Z, Mo P, Li X, Ma Z, Song S, et al. Clinical characteristics and outcomes of older patients with coronavirus disease 2019 (COVID-19) in Wuhan, China (2019): a single-centered, retrospective study. J Gerontol A Biol Sci Med Sci 2020.

[16] Chuanyu Hu ZL, Yanfeng Jiang, Xin Zhang, Oumin Shi, Kelin Xu, Chen Suo, Qin Wang, Yujing Song, Kangkang Yu, Xianhua Mao, Xuefu Wu, Mingshan Wu, Tingting Shi, Wei Jiang, Lina, Damien C Tully Lei Xu, Li Jin, Shusheng Li, Xuejin Tao, Tiejun Zhang, Xingdong Chen. Early prediction of mortality risk among severe COVID-19 patients using machine learning. 2020:2020.2004.2013.20064329.

[17] Deng Y, Liu W, Liu K, Fang YY, Shang J, Zhou L, et al. Clinical characteristics of fatal and recovered cases of coronavirus disease 2019 (COVID-19) in Wuhan, China: a retrospective study. Chin Med J (Engl) 2020.

[18] Du RH, Liang LR, Yang CQ, Wang W, Cao TZ, Li M, et al. Predictors of Mortality for Patients with COVID-19 Pneumonia Caused by SARS-CoV-2: A Prospective Cohort Study. Eur Respir J 2020.

[19] Fu Y-q, Sun Y-l, Lu S-w, Yang Y, Wang Y, Xu F. Impact of blood analysis and immune function on the prognosis of patients with COVID-19. medRxiv 2020:2020.2004.2016.20067587.

[20] Li K, Chen D, Chen S, Feng Y, Chang C, Wang Z, et al. Radiographic Findings and other Predictors in Adults with Covid-19. 2020.

[21] Luo X, Xia H, Yang W, Wang B, Guo T, Xiong J, et al. Characteristics of patients with COVID-19 during epidemic ongoing outbreak in Wuhan, China. 2020:2020.2003.2019.20033175.

[22] Ruan Q, Yang K, Wang W, Jiang L, Song J. Clinical predictors of mortality due to COVID-19 based on an analysis of data of 150 patients from Wuhan, China. Intensive Care Med 2020.

[23] Shi Q, Zhao K, Yu J, Jiang F, Feng J, Zhao K, et al. Clinical characteristics of 101 non-surviving hospitalized patients with COVID-19: A single center, retrospective study. medRxiv 2020:2020.2003.2004.20031039.

[24] Wang L, He W, Yu X, Hu D, Bao M, Liu H, et al. Coronavirus disease 2019 in elderly patients: Characteristics and prognostic factors based on 4-week follow-up. J Infect 2020.

[25] Wang Y, Lu X, Chen H, Chen T, Su N, Huang F, et al. Clinical Course and Outcomes of 344 Intensive Care Patients with COVID-19. Am J Respir Crit Care Med 2020.

[26] Xu Y. Dynamic profile of severe or critical COVID-19 cases. medRxiv 2020.

[27] Yang X, Yu Y, Xu J, Shu H, Xia J, Liu H, et al. Clinical course and outcomes of critically ill patients with SARS-CoV-2 pneumonia in Wuhan, China: a single-centered, retrospective, observational study. Lancet Respir Med 2020.

[28] Zhang F, Yang D, Li J, Gao P, Chen T, Cheng Z, et al. Myocardial injury is associated with in-hospital mortality of confirmed or suspected COVID-19 in Wuhan, China: A single center retrospective cohort study. medRxiv 2020:2020.2003.2021.20040121

[29] Zhou F, Yu T, Du R, Fan G, Liu Y, Liu Z, et al. Clinical course and risk factors for mortality of adult inpatients with COVID-19 in Wuhan, China: a retrospective cohort study. Lancet 2020;395:1054–1062.

[30] Chinese Digestion Association Chinese Medical Doctor Association, Chinese Society of Hepatology Chinese Medical Association. The protocol for prevention, diagnosis and treatment of liver injury in coronavirus disease 2019. Zhonghua Gan Zang Bing Za Zhi 2020;28:217–221.

[31] Hu LL, Wang WJ, Zhu QJ, Yang L. Novel coronavirus pneumonia related liver injury: etiological analysis and treatment strategy. Zhonghua Gan Zang Bing Za Zhi 2020;28:E001.

[32] Iba T, Levy JH. Inflammation and thrombosis: roles of neutrophils, platelets and endothelial cells and their interactions in thrombus formation during sepsis. J Thromb Haemost 2018;16:231–241.

[33] Levi M, van der Poll T. Coagulation and sepsis. Thromb Res 2017;149:38–44.

